# Sleep disruption after brain injury is associated with worse motor outcomes and slower functional recovery

**DOI:** 10.1101/2020.02.16.20022384

**Authors:** Melanie K Fleming, Tom Smejka, David Henderson Slater, Veerle van Gils, Emma Garratt, Ece Yilmaz Kara, Heidi Johansen-Berg

## Abstract

**Background and Aims:** Sleep is important for consolidation of motor learning, but brain injury may affect sleep continuity and therefore rehabilitation outcomes. This study aims to assess the relationship between sleep quality and motor recovery in brain injury patients receiving inpatient rehabilitation.

**Methods:** 59 patients with brain injury were recruited from two specialist inpatient rehabilitation units. Sleep quality was assessed (up to 3 times) objectively using actigraphy (7 nights) and subjectively using the Sleep Condition Indicator. Motor outcome assessments included: Action Research Arm test (upper limb function), Fugl Meyer assessment (motor impairment) and the Rivermead Mobility Index. The functional independence measure (FIM) was assessed at admission and discharge by the clinical team. 55 age and gender matched healthy controls completed one assessment.

**Results:** Inpatients demonstrated lower self-reported sleep quality (p<0.001) and more fragmented sleep (p<0.001) than controls. For inpatients, sleep fragmentation explained significant additional variance in motor outcomes, over and above that explained by admission FIM score (p<0.017), such that more disrupted sleep was associated with poorer motor outcomes. Using stepwise linear regression, sleep fragmentation was the only variable found to explain variance in rate of change in FIM (R^2^_adj_ = 0.12, p = 0.03), whereby more disrupted sleep was associated with slower recovery.

**Conclusions:** Inpatients with brain injury demonstrate impaired sleep quality, and this is associated with poorer motor outcomes and slower functional recovery. Further investigation is needed to determine how sleep quality can be improved and whether this affects outcome.

## Introduction

Sleep disturbance is a common complaint after brain injury, including stroke, with a high proportion (30-70%) of patients presenting with impaired subjective sleep quality and meeting the criteria for at least one sleep disorder ^1–4^. When compared to controls, reduced sleep efficiency, increased wake time, reduced rapid eye movement (REM) sleep and a lower ratio of non-REM sleep to wakefulness have all been demonstrated using polysomnography (PSG) ^5–7^. Sleep disturbance could be a result of direct damage to brain areas, or due to secondary effects such as being in the hospital environment, depression, anxiety or pain, and could potentially impact on rehabilitation through reduced engagement or impaired learning and consolidation ^8^.

There is some evidence for improvements in sleep quality from the acute to the chronic stage of stroke ^6,9^, however stroke survivors at the chronic stage continue to have impaired subjective and objective sleep quality and worse quality of life than controls ^10,11^. Interestingly, the longer the time since stroke, the worse the perceived daytime sleepiness becomes ^12^. This suggests that sleep disturbance may be persistent throughout the rehabilitation period for some patients, and changes within this time frame in patients with different types of brain injuries are yet to be determined.

The link between sleep quality and function after stroke and brain injury is currently emerging. Siccoli et al ^13^ demonstrated a cross-sectional correlation between the National Institute for Health Stroke Scale (NIHSS) score and wake after sleep onset (WASO), using PSG in the acute stage post-stroke, but their sample was small (11 patients). A larger study^14^ found a cross-sectional relationship between subjective sleep quality and the functional ambulation score after stroke, but had no objective measures of sleep quality. Similarly, Kalmbach et al ^15^ found that patients with subjective difficulties initiating sleep had lower function at multiple time-points over the first 6 months of recovery from traumatic brain injury (TBI). Sleep variables, such as total sleep time, WASO and daytime napping, have also been shown to explain significant variance in Barthel Index (BI) score at the acute stage of stroke ^16,17^, and the percentage of sleep stages I and REM are negatively associated with NIHSS ^7^.

There is evidence to suggest that the relationship between sleep quality and function may be bi-directional. Kalmbach et al ^15^ found that poorer subjective sleep quality at one time-point in the recovery period after TBI was associated with more functional impairment at the next assessment. Given that the opposite was also found (greater functional impairment at one time-point was associated with poorer sleep at the next assessment), it may be that function can impact on sleep quality which in turn may impact on recovery.

However, there is little research to indicate whether sleep quality over the rehabilitation period correlates with outcome or change in function over time, and studies that are available are somewhat inconsistent in their findings. Iddagoda et al ^4^ reported that the change in subjective sleep quality from pre-stroke to discharge from hospital correlated with the change in the functional independence measure (FIM). However, they assessed “pre-stroke” sleep quality on admission, so it is not clear how reliable their reporting would be at this time and they did not assess sleep quality at any other time during their stay other than at discharge. Nevertheless, this is consistent with a study finding that the presence of sleep disordered breathing at the acute stage is associated with reduced modified Rankin scale (mRS) and BI at 6 weeks post-stroke ^18^ and studies have demonstrated that stroke patients with poor functional outcome (mRS >2, Canadian Neurological score (CNS) < 6.5 or BI < 90)) have a lower sleep efficiency, less REM sleep or a reduced REM sleep latency at the acute stage than those with a better outcome ^7,19,20^.

In contrast, Joa et al ^21^ found no difference in the change in NIHSS or BI between patients reporting sleep disturbance at 1 month post-stroke and those reporting no disturbance. They did, however, find that the group reporting no sleep disturbance had a greater improvement in the Berg Balance scale (BBS). This was particularly evident for the moderate-severe stroke patients compared with mild (on the basis of NIHSS score at 1 week post-stroke), suggesting sleep may have a greater impact on functional recovery in those who have the most re-learning to achieve. The studies by Iddagoda et al ^4^ and Joa et al ^21^ used only subjective sleep measures and many of the studies have divided participants into groups based on outcome or the presence/absence of sleep disturbance, rather than examining both sleep quality and outcome as a continuum which may be more sensitive to differences across participants. Bakken et al ^17^ did assess objective sleep quality as a continuum, using actigraphy, but demonstrated no correlation between sleep variables in the acute stage and BI at 6 months post-stroke. In contrast, Vock et al ^9^ found, using PSG, that higher WASO or lower sleep efficiency at the acute stage post-stroke was associated with worse outcome (mRS score) at discharge. Similarly, Huang et al ^16^ demonstrate, using PSG, that total sleep time correlates positively, and sleep latency correlates negatively, with the change in BI with rehabilitation. The authors present a model including hypertension, sleep latency, percentage of time in stage N1 of sleep and desaturation, which explains 25% of the variance in the change in BI.

As there is no clear consensus on the relationship between sleep quality measures and the rate of recovery with rehabilitation, and it is unclear how sleep quality changes over the course of rehabilitation, we sought to conduct a prospective assessment of sleep quality in neurological inpatients and explore the relationship with neurorehabilitation outcomes. We therefore assessed objective and subjective sleep quality at up to three time-points throughout the rehabilitation period, and examined the relationship between sleep quality and different rehabilitation outcome measures in patients with moderate to severe brain injury. Specifically, we aimed to address the following questions:

1. Does sleep quality at a single time-point correlate with function/impairment at that time-point?
2. Does sleep quality change over the inpatient rehabilitation period?
3. Does objective sleep quality averaged over the inpatient rehabilitation period explain variance in motor outcomes over that explained by baseline function?
4. Does objective or subjective sleep quality averaged over the inpatient rehabilitation period explain variance in the rate of recovery over that explained by covariates such as initial independence, age and time since injury?

## Methods

### Participants

This was a prospective observational study, based in the Oxford Centre for Enablement Neurological Rehabilitation Unit (Oxford University Hospitals NHS Foundation Trust; June 2017-October 2019) and the Oxfordshire Stroke Rehabilitation Unit (Oxford Health NHS Foundation Trust; May 2019 - October 2019). Potential participants were screened for eligibility by the clinical and research teams following admission and approximately weekly thereafter. Inclusion criteria were: acquired brain injury with definitive onset (stroke, traumatic brain injury, haemorrhage, hypoxic brain injury) requiring motor rehabilitation (upper and/or lower limb). Exclusion criteria were: inability to provide informed consent, other neurological or psychiatric conditions, pre-existing sleep disorder. Patients with aphasia or cognitive impairment limiting the ability to provide informed consent were considered weekly and approached only if sufficient improvement was made throughout their stay to enable informed consent. This judgement was made by the multidisciplinary clinical team (including doctors, speech and language therapists and psychologists). The study was approved by the National Research Ethics Service (11/H0605/12) and all participants provided written informed consent.

In total, 197 patients were screened between June 2017 and October 2019 (Figure 1). Of these, 121 were found to be ineligible, 14 declined to participate and 62 provided written informed consent. Of those providing consent, 3 withdrew without any usable data, leaving 59 for analysis. Diagnoses included traumatic brain injury (TBI, n=9), ischaemic stroke (STROKE, n=30), intracerebral haemorrhage (ICH, n=10), subarachnoid haemorrhage (SAH, n=4) and other brain injury (OTHER, n=6). Patients were admitted to the units at variable times after their brain injury (median 29 days, range 3 – 247).

**Fig. 1.**
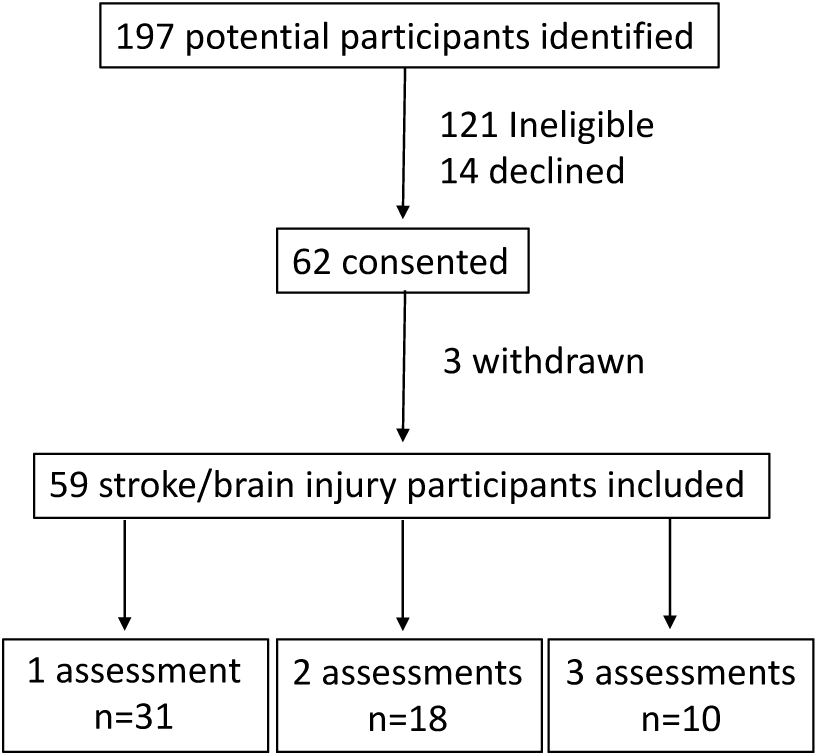
Recruitment flowchart.

Where possible, sleep quality and motor assessments were conducted as soon as possible after admission (EARLY; mean 22 days post-admission), at mid-point of their stay (MID; mean 59 days) and prior to discharge (LATE; mean 91 days). However, in some cases only one or two assessments were possible due to delays in recruitment after admission, short admissions or unexpected early discharge (Fig 1).

Additionally, sleep quality was assessed at one time point for a cohort of 55 age (mean (SD): 56.5 (17.6)) and gender (27 male, 28 female) matched, community dwelling, healthy controls in order to confirm that this cohort of inpatients demonstrated impaired sleep quality.

### Assessments

Sleep quality was assessed through actigraphy (Motionwatch, Camntech Ltd, Cambridge, UK), and the sleep condition indicator (SCI) ^22^. A motionwatch was placed on each wrist and worn continuously for 7 days and nights at each assessment timepoint. The actigraph can be used to predict when the body is in periods of sleep in comparison to wake under the assumption of the body being motionless during deep sleep. Therefore, parameters such as sleep fragmentation can be calculated ^23^ in the hospital environment. During this time, participants also completed a sleep diary to indicate what time they tried to go to sleep and what time they woke up each day. They also indicated on a 5-point scale how well they slept each night (very poor, poor, fair, good or very good). If they were unable to complete the sleep diary then the researchers or therapists assisted them to do so or approximate sleep/wake times were taken by the research team from clinical notes. The sleep condition indicator (SCI) was completed during the 7-day period (at the same time as the motor assessments) to provide a self-report measure of sleep quality and the impact of sleep disturbance on daytime function. Additionally, anxiety and depressive symptoms were assessed using the hospital anxiety and depression scale (HADS) during the 7-day sleep monitoring period.

Motor assessments were also conducted by a researcher (MKF) during the 7-day sleep monitoring period. These included the action research arm test (ARAT; max score 57) ^24^ to assess function of the upper limb, and the Fugl-Meyer scale (FMA; max score 100) ^25^ to assess upper and lower limb motor impairment.

The Functional Independence Measure^26^ (max score 126) and the Barthel Index^27,28^ (max score 20) were used to measure function and independence in activities of daily living at admission and discharge. These measures are scored by the clinical team as a matter of clinical practice. Additionally, the Rivermead mobility index^29^ (RMI; max score 15) score obtained routinely by the clinical team for some patients was recorded where possible. The clinical team had no access to sleep quality measures of any of the participants.

### Analysis

Based on a previous study, demonstrating a correlation between WASO early after stroke and BI at discharge ^9^, we estimated a likely correlation of 0.45, and therefore a sample size of 36 participants would be required (α=0.05 and 80% power). However, as we sought to perform stepwise linear regression we aimed to recruit a higher sample if possible over the 21 months.

Measures of sleep quality were taken from the Motionwatch attached to the wrist of the less-affected arm for inpatients or the non-dominant arm of healthy controls, using Motionware software (Camntech Ltd). Data were averaged by the monitor into 30 s epochs. Sleep measures included: assumed sleep (hours, minutes); actual sleep time (hours, minutes); WASO (hours, minutes), and the fragmentation index.

Statistical analysis was performed with SPSS version 25 (IBM inc.) and Graphpad Prism 8.3.0 (GraphPad Software, San Diego, California, USA). Differences in characteristics and sleep quality between patients and controls were assessed using independent samples t-tests, or Mann Whitney U tests as appropriate, with effect sizes (Cohen’s d) reported where significant differences were found. Group characteristics based on number of assessment sessions or diagnosis group were compared using one-way ANOVA, Kruskal Wallis or Chi square tests as appropriate. Where group differences were significant *post hoc* comparisons were conducted with Bonferroni correction for multiple comparisons.

Cross-sectional relationships between sleep quality, motor function/impairment and functional independence were assessed using data obtained from the first assessment only. Spearman correlations were conducted between objective sleep quality measures; WASO and fragmentation index, and subjective sleep quality (SCI score) with ARAT and FMA at the first assessment and FIM at admission. An adjusted significance level of p < 0.005 was used to compensate for multiple correlations.

Changes in sleep quality (WASO, fragmentation index, SCI score) over the rehabilitation period were assessed for participants with two or three assessments, using a linear mixed model with assessment time (EARLY, MID, DISCHARGE) as the fixed effect and participant as the random effect.

We also sought to determine whether objective sleep quality (WASO, fragmentation index) averaged over the rehabilitation period explained variance in motor function (ARAT, FMA, RMI) at discharge from the rehabilitation unit, over and above baseline severity of injury (assessed as baseline FIM). To ensure that outliers did not influence the model, data greater than 2 standard deviations from the mean were removed prior to analysis (max 4 data-points in any one measure). A hierarchical regression analysis was used. Initially, FIM at admission was entered alone into the regression model to determine the proportion of variance explained by baseline severity. Then, either WASO and fragmentation index were added using stepwise selection, to determine whether these variables increased the variance explained. An adjusted significance of p < 0.017 was used to compensate for three regression models.

Finally, we wanted to determine whether any sleep or demographic factors could explain variance in the rate of recovery of functional independence (FIM; calculated as (discharge-admission)/length of stay in days). To ensure that outliers did not influence the model, data greater than 2 standard deviations from the mean were removed prior to analysis (max 4 data-points in any one measure). Stepwise linear regression was conducted with the dependent variable of rate of change in FIM and independent variables of sleep quality (WASO, fragmentation index, SCI score), average HADS score over the rehabilitation period, age, Barthel index score (BI) at admission and time since injury at admission. Pairwise deletion was utilised to enable associations between variables to be calculated in the case of missing data in one variable.

## Results

### Inpatient Characteristics

There were no differences in age (F_2,56_ = 2.365, p = 0.103), sex (χ^2^(2)=0.320, p = 0.852), days since injury at admission to the rehabilitation unit (χ^2^(2)=3.958, p = 0.138), days since admission at recruitment (χ^2^(2)=3.840, p = 0.147), diagnosis (χ^2^(8)=8.873, p = 0.353), BI at admission (χ^2^(2)=3.016, p = 0.221) or FIM at admission (F_2,56_ = 0.346, p = 0.709) between those with 1, 2 or 3 assessments completed. There was a difference in the length of stay (χ^2^(2) = 16.657, p < 0.001), such that those with 1 assessment had a shorter length of stay than those with 2 or 3 assessments.

Participants characteristics for each diagnosis group are presented in Table 1. There were no differences in age (F_4,54_ = 1.728, p = 0.157), FIM at admission (F_4,54_ = 2.045, p = 0.101), BI at admission (χ^2^(4) = 2.254, p = 0.689), length of stay (χ^2^(4) = 3.056, p = 0.549), first assessment WASO (χ^2^(4) = 4.771, p = 0.312), fragmentation (χ^2^(4) = 8.237, p = 0.083) or SCI (χ^2^(4) = 7.331, p = 0.119) between the different diagnosis groups. There was a significant difference for time since injury at admission (χ^2^(4) = 16.865, p = 0.002). *Post hoc* Mann Whitney U Tests found that those with ICH were admitted to the rehabilitation unit more quickly than either TBI (U=9.5,0 = 0.004) or SAH (U=0.0, p = 0.005).

**Table 1.** Inpatient characteristics

|  | **STROKE** | **TBI** | **ICH** | **SAH** | **OTHER** | **All** |
| --- | --- | --- | --- | --- | --- | --- |
| **n** | 30 | 9 | 10 | 4 | 6 | 59 |
| **Age (years)** |  |  |  |  |  |  |
| *Mean (SD)* | 61 (20) | 49 (20) | 63 (19) | 56 (13) | 43 (21) | 57 (20) |
| **Sex** |  |  |  |  |  |  |
| *Male:Female* | 14:16 | 8:1 | 7:3 | 4:0 | 5:1 | 38:21 |
| **Time since injury at admission (days)** |  |  |  |  |  |  |
|  | 22<br>(6-246) | 43<br>(23-108)<sup>^</sup> | 14<br>(3-40) | 75<br>(46-247)<sup>^</sup> | 43<br>(20-98) | 29<br>(3-247) |
| **Time since admission at first assessment (days)** |  |  |  |  |  |  |
|  | 50<br>(19-262) | 74<br>(39-206) | 46<br>(19-108) | 101<br>(64-421) | 83<br>(40-191) | 62<br>(19-421) |
| **Length of stay (days)** |  |  |  |  |  |  |
|  | 78<br>(20-169) | 112<br>(14-146) | 54.5<br>(13-101) | 68<br>(33-181) | 98<br>(33-125) | 71<br>(13-181) |
| **SCI** |  |  |  |  |  |  |
|  | 21<br>(4-30) | 26<br>(12-30) | 13<br>(2-24) | 14<br>(9-24) | 16<br>(10-32) | 21<br>(2-32) |
| **WASO (mins)** |  |  |  |  |  |  |
|  | 84<br>(22-298) | 87<br>(7-173) | 68<br>(33-247) | 58<br>(49-81) | 66<br>(50-99) | 76<br>(7-198) |
| **Fragmentation Index** |  |  |  |  |  |  |
|  | 47<br>(25-131) | 29<br>(14-68) | 39<br>(26-96) | 41<br>(20-44) | 33<br>(19-65) | 42<br>(14-131) |
| **Admission scores** |  |  |  |  |  |  |
| FIM | 62<br>(30-101) | 48<br>(9-78) | 66<br>(19-83) | 66<br>(19-83) | 78.5<br>(52-92) | 61<br>(19-114)* |
| BI | 6.5<br>(2-15) | 6<br>(3-8) | 6.7<br>(1-16) | 8<br>(2-14) | 10<br>(2-17) | 7<br>(1-17)* |
| **Discharge scores** |  |  |  |  |  |  |
| FIM | 93.5<br>(48-123) | 100<br>(41-120) | 86.5<br>(53-120) | 101.5<br>(49-119) | 103<br>(74-123) | 97<br>(41-123) |
| BI | 13<br>(3-20) | 15<br>(7-20) | 13.5<br>(4-20) | 16.5<br>(4-20) | 16.5<br>(4-20) | 14<br>(3-20) |

Values are Median (range) unless otherwise specified. <sup>^</sup> significantly greater than for ICH group. \* significant improvement from admission to discharge

At the first assessment, there was significantly more movement of the less-affected arm per 24 hour period than the more-affected arm (Median (interquartile range) ‘motionwatch units’ – affected: 7 (3.5-26), less-affected: 40 (27.2-60.5), Z = 5.784, p < 0.001), suggesting overall less movement of the more impaired arm.

There was a significant overall group improvement in FIM (Z = 6.681, p < 0.001, d=1.56) and BI (Z = 6.630, p < 0.001, d = 1.54) scores from admission to discharge suggesting functional recovery over the rehabilitation period (Table 1).

### Inpatients demonstrate poor sleep quality

Initially we sought to confirm whether our inpatient cohort experienced poorer sleep than age-matched, community dwelling healthy controls (Fig 2). There was no difference between groups for age (t(112) = 0.262, p = 0.794), or sex (χ^2^(1)=2.725, p 0.099). There was significantly higher assumed sleep duration for inpatients (t(96.8) = 5.957, p < 0.001, d =), but actual sleep duration did not differ significantly with the Bonferroni correction (t(88.6) = 2.396, p = 0.019). Inpatients were found to have more fragmented sleep (Z = -5.336, p < 0.001, d = 1.15) and a higher WASO (Z = -4.977, p < 0.001, d = 1.05), as well as poorer subjective sleep quality (SCI; Z = 3.497, p < 0.001, d = 0.11) compared to controls. Factors which may influence sleep quality were also found to differ between groups; Inpatients had higher anxiety/depression (HADS; Z = -3.003, p = 0.003, d = 0.67) and more sedentary time (t(95.1) = 4.780, p < 0.001, d = 0.92) than controls. We therefore sought to examine whether these potential explanatory variables correlated with sleep quality for the inpatients only. HADS score was found to negatively correlate with SCI score (r=-0.474, p = 0.003), such that more anxiety/depression was associated with poorer subjective sleep quality, but not with objective sleep quality (Fragmentation r = 0.178, p = 0.291, WASO r = -0.053, p = 0.757). Sedentary time did not correlate significantly with any of the sleep quality measures with the Bonferroni correction (SCI: r = 0.089, p = 0.531, Fragmentation: r = -0.017, p = 0.903, WASO: r = -0.300, p = 0.026).

**Fig 2.**
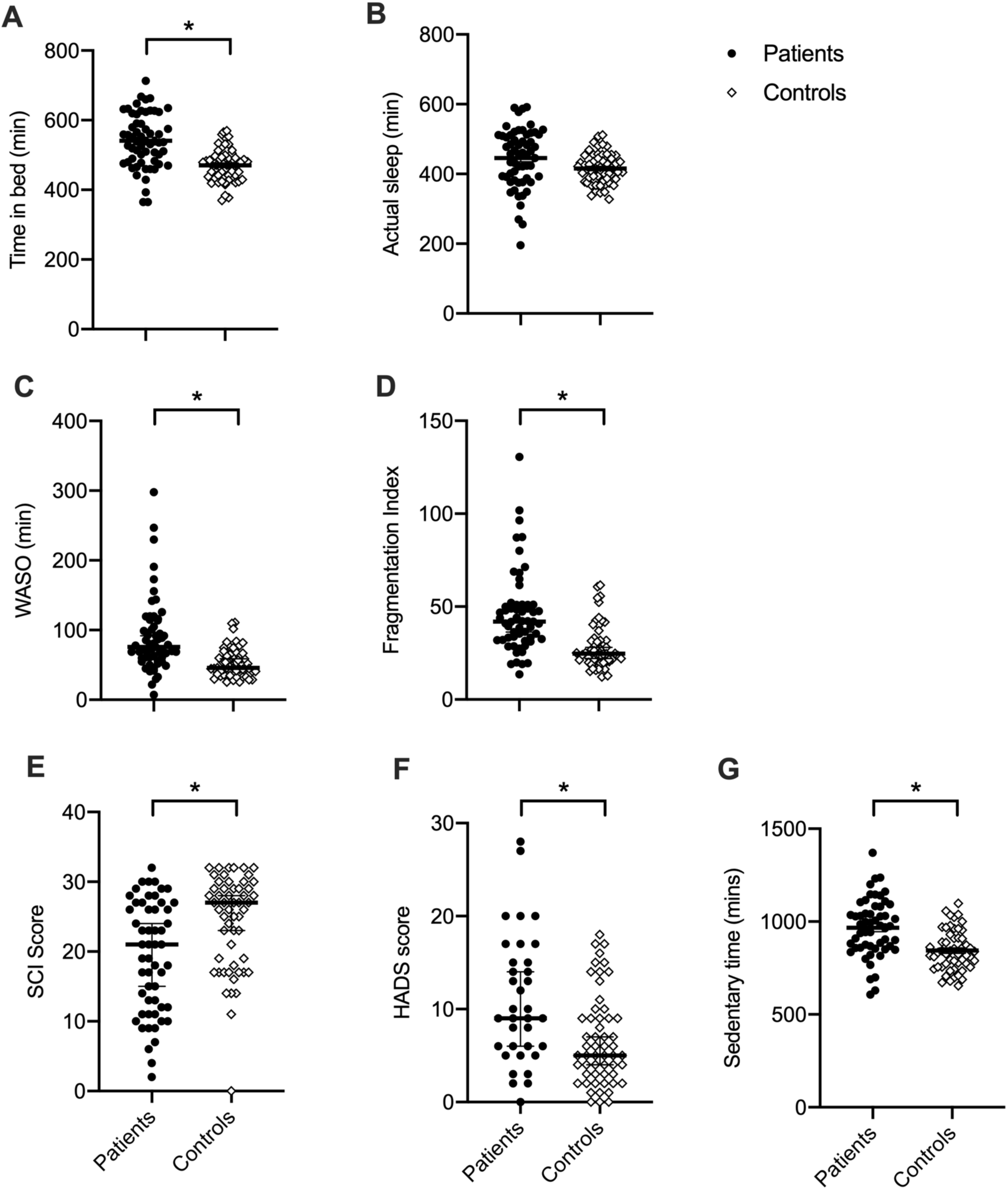
Differences between inpatients and controls. A: Inpatients show longer time in bed trying to sleep, B: Actual time asleep does not differ significantly between groups, C: Inpatients have a higher wake after sleep onset, D: Inpatients show more fragmented sleep, E: Subjective sleep quality (sleep condition indicator score) is lower for inpatients, F: Inpatients show significantly higher self-reported levels of anxiety and depression (Hospital anxiety and depression score), G: Inpatients have significantly more sedentary time per 24 hour period. Black circles = patients, Open diamonds = controls. Individual data points are shown with median or mean (black line) and standard error or the mean or 95% confidence interval as appropriate. * Mann Whitney U Test or independent samples t-test, p < 0.008.

### Cross-sectional relationships between sleep quality and function for inpatients

We then sought to assess, for inpatients, whether sleep quality at the first assessment was dependent on severity, assessed as FIM at admission and ARAT or FMA at the first assessment (Fig 3).

**Fig 3.**
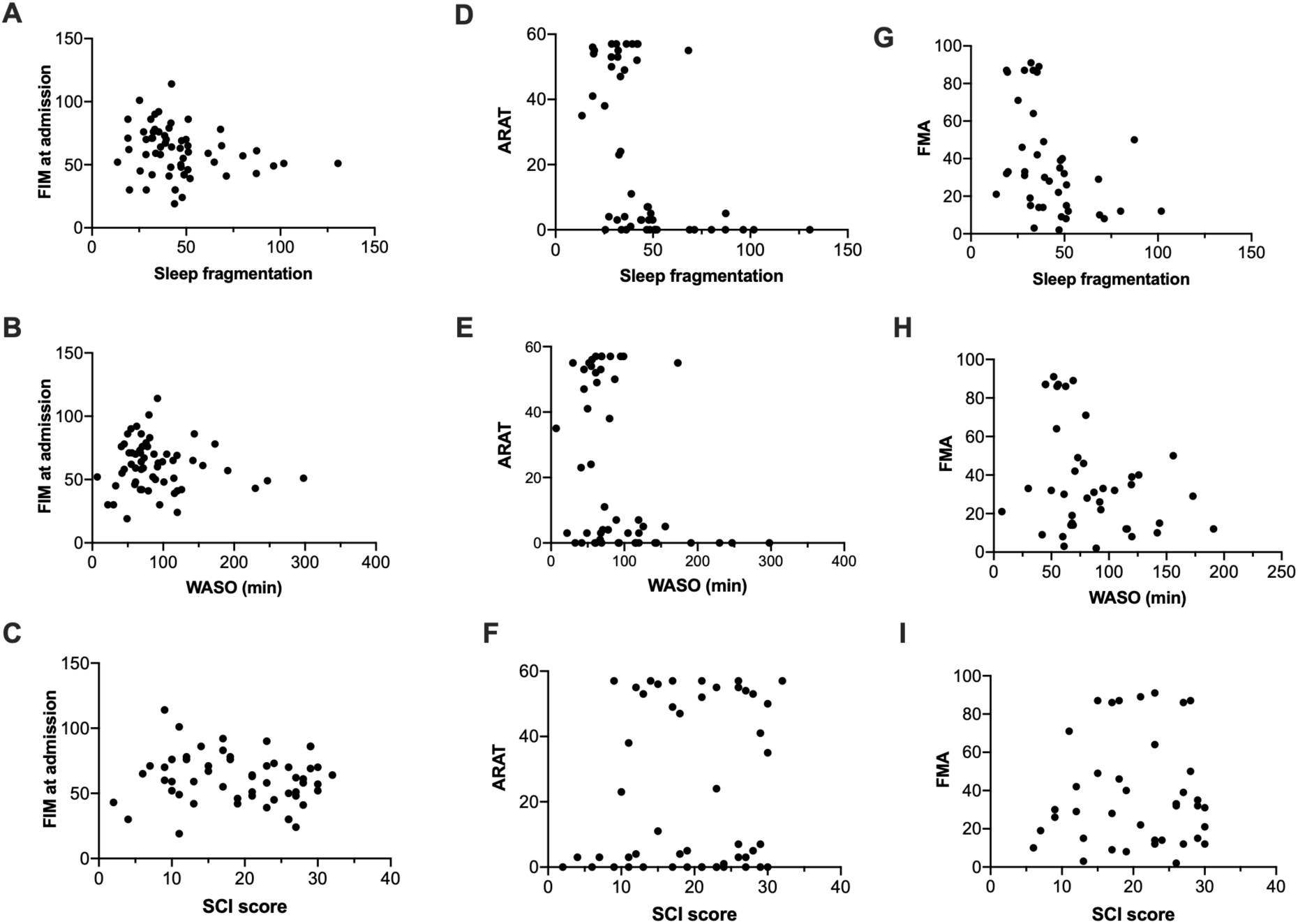
Cross sectional correlations between sleep quality measures and functional independence at admission, motor function and impairment at the first assessment for all inpatients. FIM = functional independence measure at admission, higher scores indicate more functional independence (less severe brain injury). ARAT = Action Research Arm Test; higher values indicate greater upper limb function. Higher sleep fragmentation and wake after sleep onset (WASO) indicate poorer sleep quality. Higher Sleep Condition Indicator (SCI) indicates better perceived sleep quality. FMA = Total Fugl Meyer score, higher scores indicate less motor impairment (upper and lower limb). Significant negative Spearman correlations were found for **(E)** and **(G)** (p < 0.008).

For FIM at admission (Fig 3A-C), there was a tendency for a negative correlation with Fragmentation index at the first assessment (r=-0.259, p = 0.048) which was not significant with correction for multiple correlations. There was no correlation between FIM at admission and either WASO (r=-0.038, p = 0.775) or SCI (r=-0.148, p = 0.280) at the first assessment.

For ARAT at first assessment (Fig 3D-F), there was a negative correlation with WASO (r=-0.577, p < 0.001), such that inpatients with greater WASO have worse (lower) ARAT scores. There was a tendency for a negative correlation between ARAT and Fragmentation Index (r=-0.312, p = 0.027) and no correlation between ARAT and SCI (r=0.106, p = 0.106).

For FMA at first assessment (Fig 3G-I), there was a negative correlation with Fragmentation Index (r=-0.484, p = 0.002), such that inpatients with more fragmented sleep have worse motor impairment (lower FMA score), but not for WASO (r=-0.256, p = 0.111) or SCI (r=0.027, p = 0.867).

### No change in sleep quality over time

For patients with two or more assessments, we wanted to determine whether there was a change in sleep quality alongside recovery. Linear mixed model analysis showed no significant effect of assessment time (EARLY, MID, DISCHARGE) on any of the sleep quality measures (Sleep fragmentation F_1.6,28.72_ = 1.693, p = 0.205; WASO F_1.7,30.8_ = 1.007, p = 0.366; SCI F_1.0,17.5_ = 1.894, p = 0.186, Fig 4)

**Fig 4.**
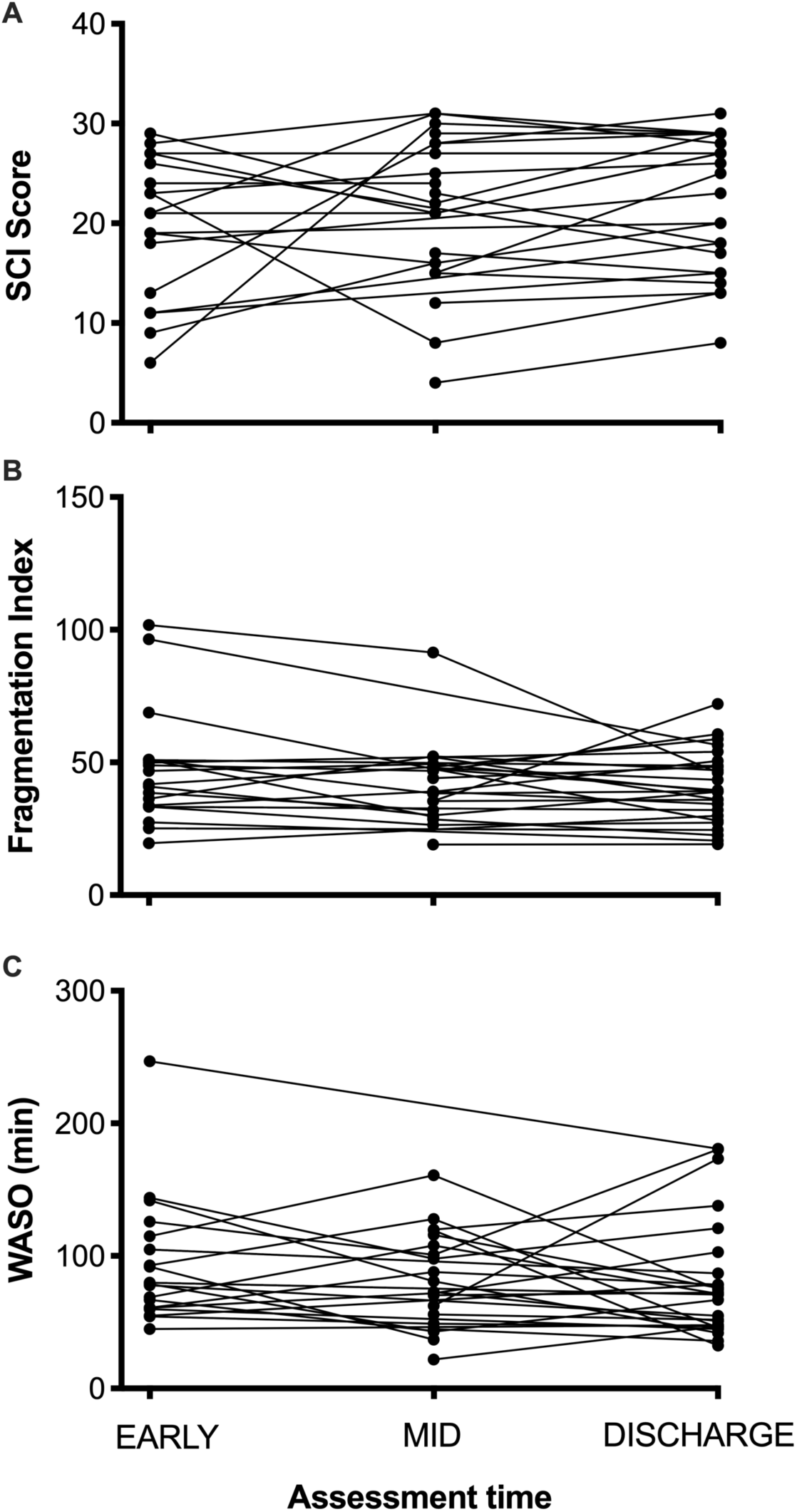
Sleep quality over each assessment for participants with two or three assessments over the inpatient stay. A: Sleep condition indicator (higher values indicate better perceived sleep quality), B: Fragmentation Index (lower values indicate better sleep quality), C: Wake after sleep onset (lower values indicate better sleep quality). There was no effect of assessment time on any of the sleep quality variables, suggesting no change in subjective or objective sleep quality over the rehabilitation period.

### Longitudinal relationships between objective sleep quality, baseline functional independence and discharge function

For ARAT at discharge, FIM at admission was not found to be a significant predictor (R_2_ = 0.132, F_1,26_ = 3.952, p = 0.057). Adding sleep fragmentation increased the variance explained to 29.2 % (ΔR^2^ = 0.160, F_1,25_ = 5.647, p = 0.025), but this did not reach significance with Bonferroni correction. WASO did not contribute to the model.

For FMA at discharge, FIM at admission was found to explain 21.1% of the variance (R^2^ = 0.211, F_1,21_ = 5.622, p = 0.031), but this was not significant with Bonferroni correction. Adding sleep fragmentation significantly increased the variance explained to 48.5 % (ΔR^2^ = 0.274, F_1,19_ = 10.1, p = 0.005), such that higher functional independence on admission and less disrupted sleep over the rehabilitation period was associated with lower motor impairment (higher FMA) at discharge. WASO did not contribute to the model.

For RMI at discharge, FIM at admission explained 18.2 % of the variance (R^2^ = 0.182, F_1,32_ = 7.115, p = 0.012). Adding sleep fragmentation significantly increased the variance explained to 43.1 % (ΔR^2^ = 0.249, F_1,31_ = 13.557, p = 0.001), such that higher functional independence at admission and less disrupted sleep over the rehabilitation period was associated with better mobility at discharge. WASO did not contribute to the model.

If a stepwise regression was used, rather than a hierarchical regression, then sleep fragmentation alone was consistently found to explain significant variance in outcome (ARAT: R^2^ = 0.212, F_1,26_ = 6.978, p = 0.014. FMA: R^2^ = 0.485, F_1,20_ = 11.188, p = 0.003. RMI: R^2^ = 0.324, F_1,32_ = 15.316, p < 0.001).

### Longitudinal relationships between sleep quality and rate of change in functional independence

Sleep fragmentation, averaged over the inpatient stay, accounted for 12 % of the variance in rate of change in FIM (R^2^_adj_ = 0.120, p = 0.030, Fig 5), such that inpatients with more fragmented sleep showed slower rates of functional recovery. SCI score, WASO or HADS score averaged over the inpatient stay, as well as age, baseline BI or time since injury at admission, did not significantly contribute to the model.

**Fig 5.**
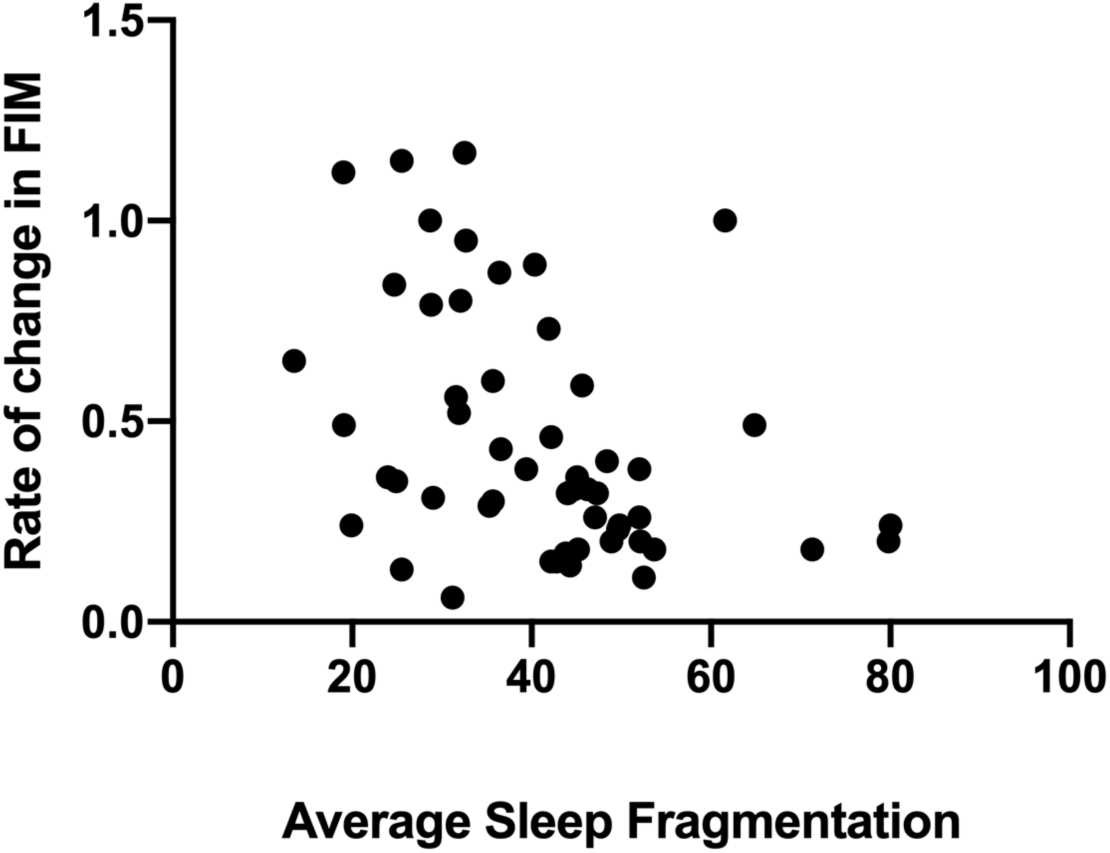
Rate of change in functional independence (FIM) as a function of sleep fragmentation averaged over the inpatient stay. FIM was found to explain 12 % of the variance in recovery.

## Discussion

This study demonstrates that people with moderate to severe brain injury, including stroke, experience significantly worse sleep quality than age-matched community dwelling healthy controls, and that more fragmented sleep is associated with poorer motor outcomes and slower recovery of functional independence throughout rehabilitation.

The inpatients demonstrated clear impairments in both objective and subjective (self-reported) sleep quality in comparison with healthy controls. This is perhaps not surprising given that hospital environments are known to be quite disruptive to sleep patterns, particularly shared rooms or units with severely disturbed patients who may call out during the night. Nevertheless, other studies have also demonstrated impairment in aspects of sleep quality in comparison with people hospitalised for non-neurological reasons ^30^, and therefore the disruption to sleep is unlikely to be solely due to the environment of the rehabilitation unit.

Sleep fragmentation may play a key role in explaining variance in outcome and recovery, as this was the only measure that was consistently found to contribute to regression models. Sleep fragmentation was consistently found to significantly increase the proportion of variance in motor outcome explained, over and above that of baseline severity (FIM at admission), indicating that patients with poor sleep demonstrate worse outcomes, even when baseline severity is taken into account. The results of the regression analysis for rate of recovery of FIM suggest that sleep fragmentation explains variance in recovery that cannot be explained by age, depression and anxiety or baseline independence in activities of daily living (BI at admission). These findings are generally consistent with previous observations that poorer functional outcome is associated with impaired sleep quality ^7,19,20^, and extends these to motor outcomes in addition to functional independence. To our knowledge this is the first study to observe significant relationships between fragmentation index and outcome/recovery assessed as a continuum rather than categorising patients as having a “good” or “poor” outcome. Further, to our knowledge this is the first study to relate sleep fragmentation averaged over the rehabilitation period to the rate of change in functional independence, suggesting that those with more disrupted sleep may recover more slowly. However, as FIM was only measured at admission and discharge, rather than multiple time-points, it was not possible to ascertain whether patients had reached a plateau in their recovery. It remains to be seen whether improving sleep quality could therefore affect rehabilitation outcomes, or whether a longer length of stay would be sufficient to improve outcome.

This study included people with a range of neurological impairments admitted to the same neurological rehabilitation units and receiving similar multidisciplinary therapy input. Unfortunately, it was not possible to gain access to clinical brain imaging to ascertain whether sleep disturbance was related to lesion extent or location. We were also not specifically aiming to assess whether sleep quality or outcome depended on the type of brain injury, though baseline comparisons suggested no clear differences in WASO, fragmentation, SCI or the time spent in rehabilitation between the brain injury subtypes. Bakken et al ^31^ similarly demonstrated that actigraphy variables (total sleep time, WASO, number of wakenings) did not differ between stroke types (ischaemic, haemorrhagic, chronic cerebral ischaemia and negative findings on CT) or between left, right and bilateral strokes. Nevertheless, it would be important for future studies to investigate whether lesion characteristics influence sleep quality or the nature of sleep disturbance.

We were surprised to find no improvement in objective or subjective sleep quality over the course of the rehabilitation period. One previous study ^6^ showed a significant improvement in WASO, sleep efficiency and apnoea-hypopnea index from acute to 3 months post stroke/TIA, but many sleep architecture measures (e.g. N1%, REM%) were unchanged and the percentage of patients with a periodic limb movement index ≥10 was actually found to get worse. Another study ^9^ found improvements in WASO and sleep efficiency from the acute to the chronic stage of stroke, although their sample at the chronic stage was just 15 patients and none of their patients were over 75 years of age which may not be particularly representative of the stroke population. Overall this finding therefore suggests that there may not be clear improvements in sleep over the early stages of recovery, and this may indicate that sleep disturbance is largely due to environmental issues, or that the neurological aspects that affect sleep quality are slow to recover.

Sleep disturbance could potentially affect rehabilitation through a reduced ability to engage in therapy activities. Worthington and Melia ^32^ report that rehabilitation unit staff feel that rehabilitation and daily activities are frequently affected for patients with acquired brain injury who demonstrate arousal disturbance. Further, more time in bed at night has been found to be associated with less daytime activity after stroke ^33^. We found significantly higher sedentary time for inpatients compared with controls, consistent with a study in chronic stroke survivors ^34^. However, in our cohort there was no correlation between sleep quality and total sedentary time, and as such there is no clear indication that those with poor sleep are engaging in rehabilitation any less than those with better sleep.

## Conclusion/Implications

Overall, this study provides evidence for a relationship between sleep fragmentation and motor outcomes as well as recovery of functional independence during neurorehabilitation. Future studies should explore factors affecting sleep quality and develop interventions to see whether sleep quality can be improved in this environment, and whether this leads to improvements in recovery.

## Data Availability

Anonymous data is available upon reasonable request.

## Acknowledgements

Thanks to all staff at the Oxford Centre for Enablement and the Oxfordshire Stroke Rehabilitation Unit for assistance with running the study. Thank you to Ximena Omlin for advice on actigraphy analysis.

This study is funded by the Wellcome Trust (Principal Research Fellowship to HJB) 110027/Z/15/Z and supported by the National Institute for Health Research (NIHR) Oxford Biomedical Research Centre. The Wellcome Centre for Integrative Neuroimaging is supported by core funding from the Wellcome Trust 203139/Z/16/Z.

## Notes

### Competing Interest Statement

The authors have declared no competing interest.

### Clinical Trial

Observational study, not a clinical trial.

